# Increased SAR-CoV-2 shedding associated with reduced disease severity despite continually emerging genetic variants

**DOI:** 10.1101/2021.02.03.21250928

**Authors:** Cynthia Y. Tang, Yang Wang, Cheng Gao, David R. Smith, Jane A. McElroy, Tao Li, Karen Segovia, Tricia Haynes, Richard Hammer, Christopher Sampson, Detlef Ritter, Christopher Schulze, Robin Trotman, Grace M Lidl, Richard Webby, Jun Hang, Xiu-Feng Wan

## Abstract

Since the first report of SARS-CoV-2 in December 2019, genetic variants have continued to emerge, complicating strategies for mitigating the disease burden of COVID-19. Positive SARS-CoV-2 nasopharyngeal swabs (n=8,735) were collected from Missouri, USA, from March-October 2020, and viral genomes (n=178) were sequenced. Hospitalization status and length of stay were extracted from medical charts of 1,335 patients and integrated with emerging genetic variants and viral shedding analyses for assessment of clinical impacts. Multiple introductions of SARS-CoV-2 into Missouri, primarily from Australia, Europe, and domestic states, were observed. Four local lineages rapidly emerged and spread across urban and rural regions in Missouri. While the majority of Missouri viruses harbored Spike-D614G mutations, a large number of unreported mutations were identified among Missouri viruses, including seven in the RNA-dependent RNA polymerase complex and Spike protein that were positively selected. A 15.6-fold increase in viral RNA levels in swab samples occurred from March to May and remained elevated. Accounting for other comorbidities, individuals test-positive for COVID-19 with high viral loads were less likely to be hospitalized (odds ratio=0.39, 95% confidence interval [CI]=0.20, 0.77) and had shorter hospital stays (hazard ratio=0.34, p=0.003) than those with low viral loads. Overall, the first eight months of the pandemic in Missouri saw multiple locally acquired mutants emerge and dominate in urban and rural locations. Although we were unable to find associations between specific variants and greater disease severity, Missouri COVID-positive individuals that presented with increased viral shedding had less severe disease by several measures.

## INTRODUCTION

Since the severe acute respiratory syndrome coronavirus 2 (SARS-CoV-2) emerged in late 2019, the virus has undergone genetic evolution. As of January 28, 2021, the United States had detected 25,301,166 cases and 423,519 deaths due to coronavirus disease 2019 (COVID-19)^1^. Emerging genetic SARS-CoV-2 variants are expected to cause prolongation of the pandemic and continued disease burdens^2^.

The massive transmission of SARS-CoV-2 and its accumulation of mutations suggests that the virus is continuing to adapt to its human host^3,4^. Over 198 sites on the viral genome containing recurrent mutations and 80 viral lineages were identified by May^5,6^. Sites on the SARS-CoV-2 genome are still undergoing positive selection^7,8^. Variants of the D614G and N501Y strains (both located on the S gene) have generated global concern for increased transmissibility with little evidence of association with disease severity^9-14^.

Despite the large number of cases, the power to detect SARS-CoV-2 variants in the United States has been insufficient. As of January 16, 2021, Missouri had experienced 452,937 confirmed cases and 6,709 deaths^15^. In this study, we investigate the emergence and spread of SARS-CoV-2 genetic variants in Central and Southwest Missouri, examine viral shedding over time, and analyze the associations among emerging genetic variants, viral shedding, and disease severity.

## METHODS

### Ethical Approval

Approved by University of Missouri Institutional Review Board (#2025449).

### Sample selection

Data from 8,735 COVID-19 positive nasopharyngeal swabs from March 7-October 31, 2020 were compiled from the Paternity Testing Corporation (PTC) Laboratories in Columbia, Missouri. Samples were collected and processed using the same protocol throughout this study, and consistently tested using the Centers for Disease Control (CDC) 2019 Novel Coronavirus Real-Time Reverse Transcriptase (RT)-PCR Diagnostic Panel^16^. Positive tests were defined as those with a threshold cycle (Ct) <45, positive nucleocapsid (N)1 and N2, and an amplified anti-human positive control. The first positive test from each individual at CoxHealth (Springfield, Missouri) or University of Missouri Health Care (UMHC; Columbia, Missouri) was used in analyses.

Viral genomes (n=184) were sequenced from swab samples and virus isolates. Available samples (92 samples from 85 patients) collected from UMHC between March 18-May 14, 2020 were sequenced. The second batch (92 samples, 90 patients) was collected between June 28-July 12, 2020 from CoxHealth after viral loads in swabs were observed to be markedly elevated. Of these samples, 136 complete viral sequences were generated with >70% coverage (eTable1). Viruses from remaining samples yielded partial genomes or had inadequate RNA quantity for sequencing.

### Chart review

Demographics, comorbidities, and hospitalization information were extracted from electronic medical records for 1,335 patients (eMethods). Ct-values were provided by PTC Laboratories. Age groups were defined in alignment with CDC reports^1^.

### Viral isolation and growth kinetics

SARS-CoV-2 viruses were recovered in Vero E6 (CRL1586™, ATCC) or Vero (NR-10385, BEI Resources) cells a maximum of three times until cytopathic effect was observed. To determine virus replication kinetics, Vero E6 cells were infected at a starting multiplicity of infection (MOI) of 0.01, and supernatants were harvested and titrated using plaque assays (eMethods).

### Genetic sequencing and assembly

SARS-CoV-2 whole genome RT-PCR amplification and next-generation sequencing was conducted using the Access Array (AA) microfluidic system (Fluidigm Corporation) and MiSeq system (Illumina)^17^. Genome assembly was constructed using Qiagen CLC Genomics Workbench 20.0.4. Amino acid variants were identified with a variant probability threshold >80% and minimum two counts, then curated manually (eMethods).

### Phylogenetic and phylogeographic analyses

To identify likely seeding viruses for Missouri outbreaks, all 110,901 SARS-CoV-2 complete genomes available on the Global Initiative on Sharing Avian Influenza Data (GISAID) consortium^18^(September 20, 2020) were downloaded. The 297 genetically closest sequences to Missouri samples were selected using an alignment-free complete composition vector algorithm^19-24^. Sequence alignments were performed using MUSCLE^25^, phylogenetic analyses using BEAST2^26,27^, positive selection analyses through PAML^28^, and sequence conservation visualization with SimPlot^29^(eMethods).

Lineages were identified by PANGOLIN v2.0.8 (github.com/cov-lineages/pangolin). Unique Missouri sub-lineages were identified when they contained at least five samples with unpublished mutations, posterior probability >0.99, and sequence identity >99%.

### Statistical analyses

Kruskal-Wallis tests were used to analyze continuous variables and Fisher exact tests for categorical variables. Logistic regression models were used to assess the effect of viral load (Ct≤20, high; Ct>20, low) on hospitalization or length of hospital stay, controlling for demographics and comorbidities. The effect of viral load on length of hospitalization was tested with a Cox Proportional Hazards survival analysis accounting for censoring due to death. In all analyses, significance was defined at alpha=0.05. Analyses were performed using SAS Studio v3.8 (Cary, Indiana)

## RESULTS

### SARS-CoV-2 introductions and outbreaks of COVID-19 in Missouri

The first known introduction of SARS-CoV-2 into Missouri was publicly announced on March 7th. Based on state data^30^, Missouri had an average of 153 (standard deviation [SD]=96) cases per week between March and May increasing to an average of 280 (SD=159) cases in June and climbed with sporadic spikes throughout the study period (Figure 1A). Meanwhile, the weekly case fatality rate peaked at 12% in early May, then progressively decreased and remained below 0.02% through October.

**Figure 1.**
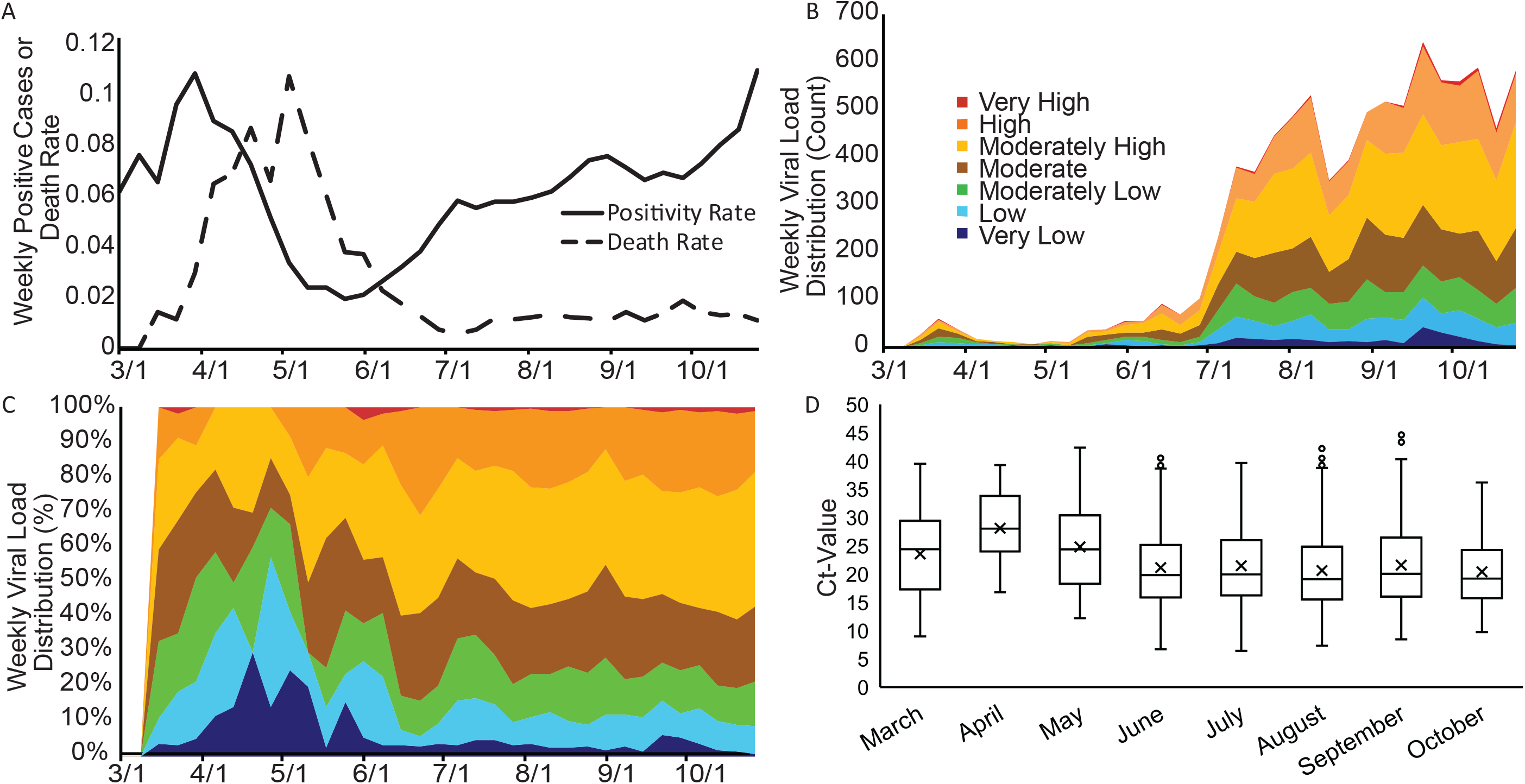
Spatiotemporal patterns of COVID-19 cases and shedding in Missouri. A) Weekly positive rate of laboratory confirmed COVID-19 infections (positive cases per test per week) and COVID-19 associated mortality (deaths per positive case per week) in Missouri; B) number of nasopharyngeal swab samples with corresponding viral load category by week in Southwest and Central Missouri; C) proportion of viral load category by week in Southwest and Central Missouri; Viral load categories: very high, Ct-value (Ct) ≤ 10; high, 10 < Ct ≤ 15; moderately high, 15 < Ct ≤ 20; moderate, 20 < Ct ≤ 25; moderately low, 25 < Ct ≤ 30; low, 30 < Ct ≤ 35; very low, Ct > 35. D) Ct-value distribution by month. Boxes represent interquartile ranges, horizontal lines within the boxes represent median Ct, and whiskers denote the minimum and maximum values. Nasopharyngeal swabs were analyzed by quantitative RT-PCR (qRT-PCR). A Ct-value of under 45 was used as a criterium for positiveness^16^.

To study whether changes in viral shedding correlates with increased positivity rates, we analyzed the cycle threshold (Ct)-values derived from quantitative RT-PCR (qRT-PCR) on nasopharyngeal swabs for 8,735 patients in Central and Southwestern Missouri between March and October (Figure 1B-D, eFigure 1). Results showed that Ct-values changed over time (P<0.0001). Viral RNA loads from March through May ranged from 8.97 to 42.33 (median=24.85). The proportion of viral load cases above moderate levels (Ct≤20) began rising in May and remained elevated through October.

### Characteristics of Study Population

We conducted chart reviews for 1,335 COVID-19 individuals from March-October 2020 (Table 1). The largest age group was 18-29 years (n=562 of 1335, 42.13%). Our dataset consisted of 55.10% female (n=735) and 44.7% male (n=596). Of hospitalized patients, there were 22 males and 22 females. The population studied consisted primarily of Caucasians (n=899, 67.39%) with 15.74% Black or African American patients (n=210). Primary comorbidities were obesity (n=599, 64.90%), diabetes (n=29, 2.17%), and hypertension (n=64, 4.80%). Overall, 44 (3.30%) patients were hospitalized with an average stay of 9.90 (SD=12.67) days. Only three of the hospitalized patients were under age 30. Four patients died.

**Table 1.** Demographic and clinical characteristics of 1,335 COVID-19 patients at initial presentation. Data reported as number (No.) of samples and percentage in parentheses of all samples within the category unless otherwise indicated. ^*^, BMI available for 923 total patients; % out of patients with available BMI.

|  | Total | Hospitalized | Not Hospitalized | Patients with Samples Submitted |
| --- | --- | --- | --- | --- |
| **Total** | 1335 | 44 | 1291 | 175 |
| **Age: Mean (SD)** | 34.35 (16.82) | 55.39 (14.61) | 33.64 (16.42) | 35.77 (16.55) |
| **Age Category: No. (%)** |  |  |  |  |
| 0-4 | 21 (1.57) | 0 | 21 (1.63) | 3 (1.71) |
| 5-17 | 89 (6.67) | 0 | 89 (6.90) | 8 (4.57) |
| 18-29 | 562 (42.13) | 3 (6.82) | 559 (43.33) | 67 (38.29) |
| 30-39 | 210 (15.74) | 5 (11.36) | 205 (15.89) | 34 (19.43) |
| 40-49 | 185 (13.87) | 5 (11.36) | 180 (13.95) | 22 (12.57) |
| 50-64 | 187 (14.02) | 19 (43.18) | 168 (13.02) | 28 (16.00) |
| 65-74 | 61 (4.57) | 8 (18.18) | 53 (4.11) | 12 (6.86) |
| 75-84 | 15 (1.12) | 4 (9.09) | 11 (0.85) | 1 (0.57) |
| >85 | 4 (0.30) | 0 | 4 (0.31) | 0 |
| **Sex: No. (%)** |  |  |  |  |
| Male | 596 (44.70) | 22 (50) | 574 (44.50) | 77 (44) |
| Female | 735 (55.10) | 22 (50) | 713 (55.30) | 98 (56) |
| Unspecified | 3 (0.20) | 0 | 3 (0.20) | 0 (0) |
| **Race: No. (%)** |  |  |  |  |
| White | 899 (67.39) | 34 (77.27) | 865 (67.05) | 150 (85.71) |
| Black or African American | 210 (15.74) | 6 (13.64) | 204 (15.81) | 13 (7.43) |
| Asian | 14 (1.05) | 0 | 14 (1.09) | 3 (1.71) |
| American Indian or Alaskan Native | 2 (0.15) | 0 | 2 (0.16) | 0 |
| Native Hawaiian or other Pacific Islander | 2 (0.15) | 0 | 2 (0.16) | 0 |
| Multi-Race | 1 (0.08) | 0 | 1 (0.08) | 1 (0.57) |
| Other Race | 53 (3.97) | 3 (6.82) | 50 (3.88) | 2 (1.14) |
| Unspecified | 153 (11.47) | 1 (2.27) | 152 (11.78) | 6 (3.43) |
| **Ethnicity: No. (%)** |  |  |  |  |
| Hispanic, Latino, Spanish Origin | 101 (7.57) | 6 (13.64) | 95 (7.36) | 16 (9.14) |
| Not Hispanic, Latino, Spanish Origin | 1101 (82.53) | 38 (86.36) | 1063 (82.40) | 153 (87.43) |
| Unspecified | 132 (9.90) | 0 | 132 (10.23) | 6 (3.43) |
| **Comorbidities: No. (%)** |  |  |  |  |
| Transplant History | 2 (0.15) | 1 (2.27) | 1 (2.27) | 1 (0.75) |
| Autoimmune Disease | 7 (0.53) | 1 (2.27) | 6 (13.64) | 8 (6.02) |
| Coagulation or Blood Disorder | 1 (0.08) | 1 (2.27) | 0 | 2 (1.50) |
| HIV/AIDS | 25 (1.87) | 4 (9.09) | 21 (1.63) | 1 (0.75) |
| Acute Kidney Injury | 1 (0.08) | 1 (2.27) | 0 | 0 |
| Chronic Kidney Disease | 4 (0.30) | 1 (2.27) | 3 (6.82) | 4 (3.01) |
| Hepatitis | 2 (0.15) | 2 (4.55) | 0 | 2 (1.50) |
| Malignancy | 3 (0.23) | 0 | 3 (6.82) | 8 (6.02) |
| Asthma | 7 (0.53) | 1 (2.27) | 6 (13.64) | 12 (9.02) |
| Diabetes | 29 (2.17) | 2 (4.55) | 27 (61.36) | 10 (7.52) |
| Chronic Obstructive Pulmonary Disease | 7 (0.53) | 4 (9.09) | 3 (6.82) | 2 (1.50) |
| Heart Disease | 2 (0.15) | 1 (2.27) | 1 (2.27) | 2 (1.50) |
| Atrial Fibrillation or Flutter | 5 (0.38) | 2 (4.55) | 3 (6.82) | 1 (0.75) |
| Hypertension | 64 (4.80) | 6 (13.64) | 58 (131.82) | 28 (2.26) |
| Heart Failure | 1 (0.08) | 1 (2.27) | 0 | 3 (2.26) |
| Obesity* | 599 (64.90) | 39 (88.64) | 560 (63.71) | 70 (67.96) |
| BMI: Mean (SD) | 28.219 (9.37) | 28.00 (9.36) | 29.411 (8.33) | 29.46 (7.69) |
| **Outcomes: No. (%)** |  |  |  |  |
| Hospitalized due to COVID-19 | 44 (3.30) | 44 (100) | 0 | 7 (6.42) |
| Length of Stay: Mean (SD) | 9.86 (12.69) | 9.86 (12.69) | NA | 11.43 (22.31) |
| Death due to COVID-19 | 4 (0.30) | 4 (9.09) | 0 | 0 |
| Ct-Value: Mean (SD) | 22.08 (7.33) | 24.07 (6.68) | 22.01 (7.34) | 18.85 (4.62) |

### Rapid evolution of COVID-19 identified through phylogenetic analyses

We collected nasopharyngeal swab samples from 175 patients for sequencing based on sample availability. 136 samples had adequate viral load for complete genome resolution (132 from clinical swabs, four from isolates) with 14 samples having only partial sequences and 38 samples where RNA quality was too low for sequencing.

Phylogenetic analysis showed that Missouri viruses encompassed eight major PANGOLIN lineages, each of which was associated with an independent introduction (Figure 2, eTable 2). The virus evolved after each initial introduction, and formed four unique Missouri sub-lineages, at least one (MO-B.1.1.b) of which circulated from the May through July collection periods. All sub-lineages were supported by posterior probabilities >0.99 and sequence identities >99.99% (Figure 2A). At least five lineages, including the four Missouri sub-lineages, co-circulated in Missouri during the week of July 2. All five lineages originated from lineage B.1 (containing D614G in the spike [S] protein), which has been predominant in Europe, Australia, and multiple states of United States (eFigure 2-3).

**Figure 2.**
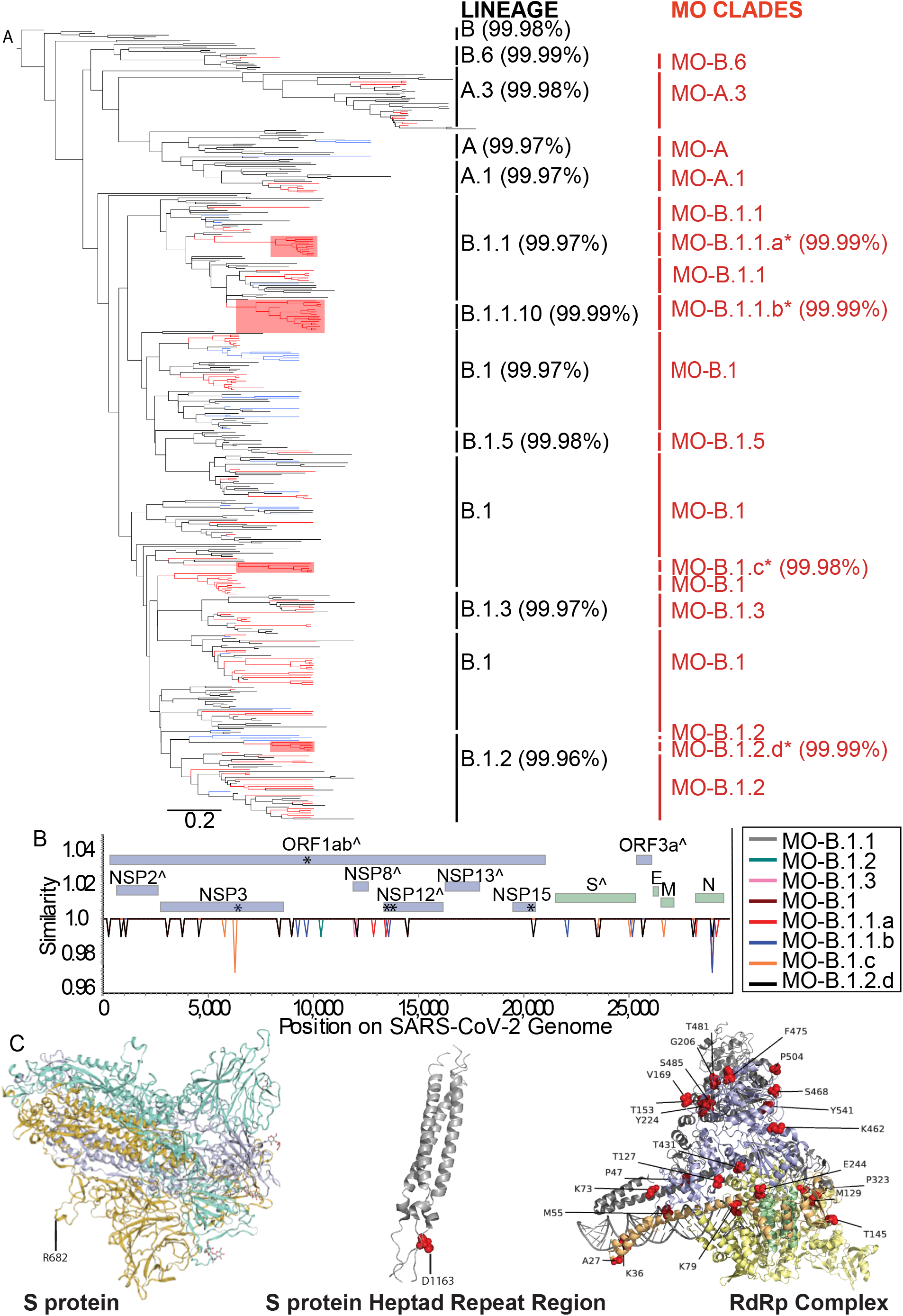
Phylogenetic analyses, genomic diversity, positive selection, and growth diversity of SARS-CoV-2 viruses in Missouri. A) Bayesian tree of SARS-CoV-2 constructed using viral genomic sequences. Sequences from nasopharyngeal swabs or isolates from this study are marked in red, Missouri samples from public databases in blue, and other original lineages sequences or closely related sequences in black. The tree was constructed using BEAST2 and rooted to hCoV-19/Wuhan/IPBCAMS-WH-01/2019. The nomenclature of genetic lineages was adapted from the PANGOLIN software (github.com/cov-lineages/pangolin); PANGOLIN lineages are labeled in black. For consistency, the Missouri specific sub-lineages, labeled in red, were also assigned based on PANGOLIN parent lineages, sequence identities, and posterior probability (see Materials and Methods). The novel lineages identified from this study are indicated by ^*^, and the viruses in these novel Missouri lineages are also shaded red in the tree. Percent sequence identities among the sequences with each lineage is shown in parentheses. Effective Sample Size (ESS) = 720. B) genomic diversity among the reference genome, Wuhan/IPBCAMS-WH-01/2019, and the SARS-CoV-2 viruses in representative Missouri lineages. Genome-wide distribution of mutations by Missouri lineage along the SARS-CoV-2 genome were analyzed by using SimPlot^29^. Green boxes represent major structural proteins. Blue boxes represent nonstructural or accessory proteins. ORF, open reading frame; NSP, nonstructural protein; S, spike; M, membrane; N, nucleocapsid; E, envelope; ^^^, protein contains sites undergoing positive selection; ^*^, sites of unique, Missouri strains in Missouri community acquired lineage defining mutations. C) Visualization of sites undergoing positive selection on S protein (Swiss-Model A0A6H1PJZ3, PDB-2FXP) and RdRp complex (PDB-6XEZ): NSP7 (green), NSP8 (orange), NSP12 (yellow), NSP13 (blue). Sites undergoing positive selection are labeled and highlighted in red.

Molecular characterization identified mutations across multiple regions of the viral genome (Figure 2B). The previously reported S-D614G and nonstructural protein [NSP]12-P314L mutations appeared in most of the Missouri samples. Compared to their precursor viruses, the Missouri viruses had 126 new mutations (eTable 3). NSP3 contained the most mutations (28 of 126 distinct mutations), followed by S (13), Nucleocapsid (N) (11), and NSP2 (11). The most common mutations include NSP12-C22F (n=18), NSP4-M366I (18), open reading frame [ORF]8-S47F (12), NSP12-A2V (11), and N-V270L (10).

Six unique mutations (Figure 2B, eFigure 2) were detected in four sub-lineages that appear to have emerged and spread in the Southwestern region of the state. MO-B.1.1.a (n=11 sequences) had mutation NSP12-A2V; 10 of these viruses were from Springfield, Missouri collected between July 2-9; MO-B.1.1.b (n=18) contained NSP4-M366I and NSP12-C22F from patients living within 60 miles of Springfield between May 14-July 9, 2020; MO-B.1.c (n=6) with NSP3-N1178T and NSP3-A1179T includes six samples with all but one arising from Monett, an urban center in Southwestern Missouri (50 miles southwest from Springfield), between July 2-6, 2020; MO-B.1.2.d (n=6) with NSP15-P262L were from Springfield or Brighton, Missouri (20 miles north of Springfield) between July 5-7, 2020.

To explore how SARS-CoV-2 viruses were adapting in Missouri, we determined variant sites undergoing selective pressure (Figure 2B-C and eTable 4). Multiple sites along the S protein, the protein mediating host receptor binding and viral entry, and the RNA-dependent RNA polymerase (RdRp) complex (NSP7, NSP8, NSF12, and NSP13) had evidence of positive selection (Figure 3B). Seven of these positively selective sites were unique to Missouri isolates (i.e., D1163Y in Spike, K36Q and T145I in NSP8, and T172I, T431I, K460R, and S468L in NSP13). NSP8 is a cofactor to NSP12 (RdRp) and is necessary for RNA synthesis, NSP12 functions in viral replication and transcription, and NSP13 works with NSP12 in replication and mRNA capping^31^.

**Figure 3.**
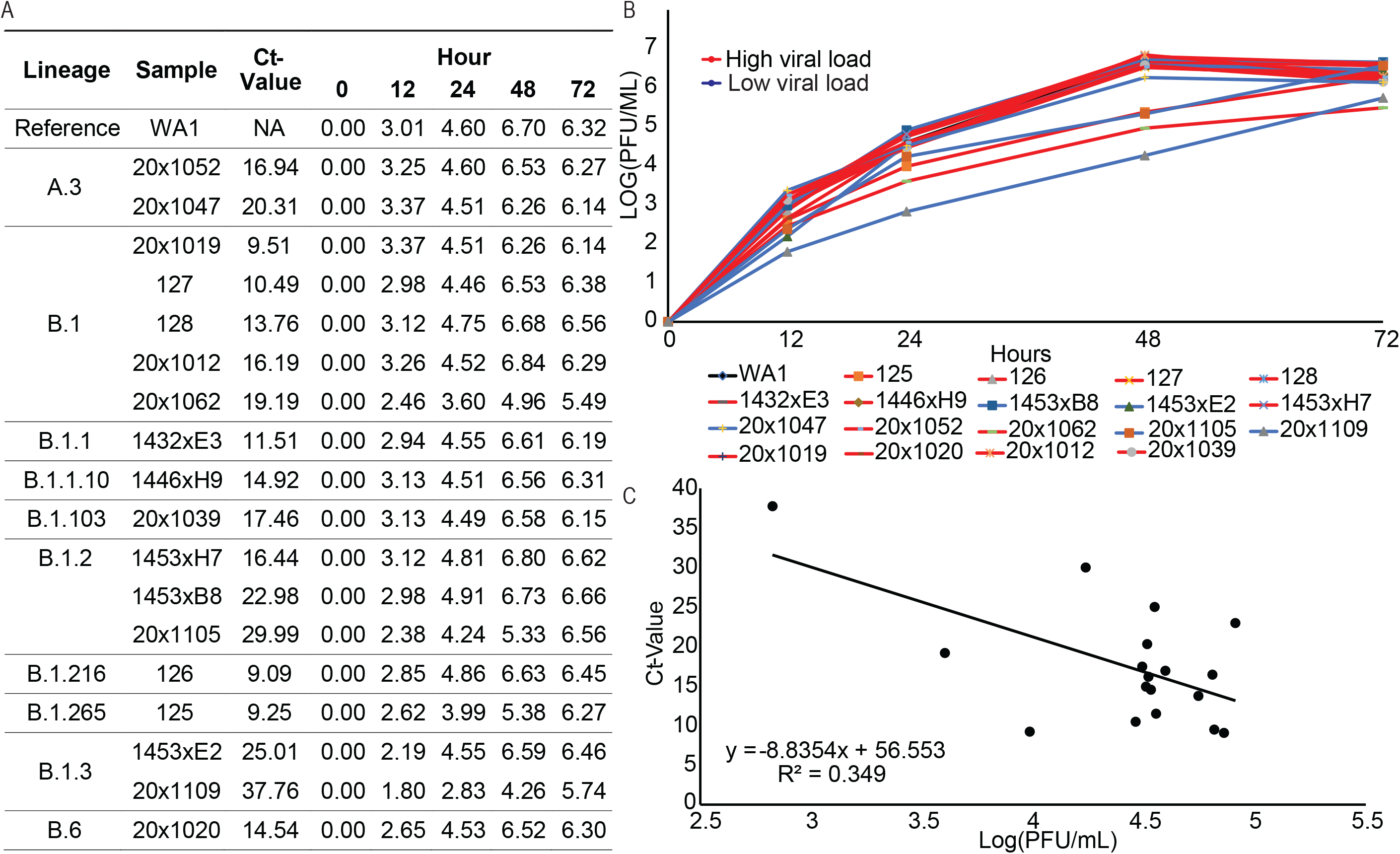
High viral loads in nasopharyngeal swabs are associated with rapid growth of the SARS-CoV-2 in cell culture. A) Representative sequences of SARS-CoV-2 strains and viral loads across Missouri lineages compared with reference sequence WA1, the original sequence from Washington state. Growth is reported as log(PFU/mL) and measurements were taken at 12, 24, 36, and 72 hours; B) Growth kinetics of the representative SARS-CoV-2 strains. High viral loads (Ct<20) are indicated with red lines and low viral loads (Ct > 20) are indicated in blue lines. WA1 is black. Marker shapes and colors differentiate the samples; C) Linear regression of Ct-values by qRT-PCR and log(PFU/mL) of the 18 representative samples. PFU, plaque forming unit.

### Re-infection with different viruses

Of the selected samples, we collected multiple samples at different time points for four patients and found one case of reinfection (eFigure 4). This patient was a female in her 20s with asthma, obesity, anxiety, and depression, who reported chills, sore throat, dizziness, rhinorrhea, and fever during her initial positive COVID-19 test in March 2020. She was discharged and instructed to self-isolate. After two weeks, her symptoms had waned to encompass only cough and fatigue, and she was tested again due to her return-to-work requirements with another positive result. Interestingly, the two samples were from two distinct SARS-CoV-2 lineages; the first sample belonged to PANGOLIN A.3 lineage, whereas the second belonged to PANGOLIN B.1.1 lineage, suggesting that the patient was reinfected by a different virus within a two-week period. Compared with those in A.3 lineage, the B.1.1. lineage virus had four new mutations ORF1ab-P4715L, S-D614G, N-R203L, and N-G204R.

**Figure 4.**
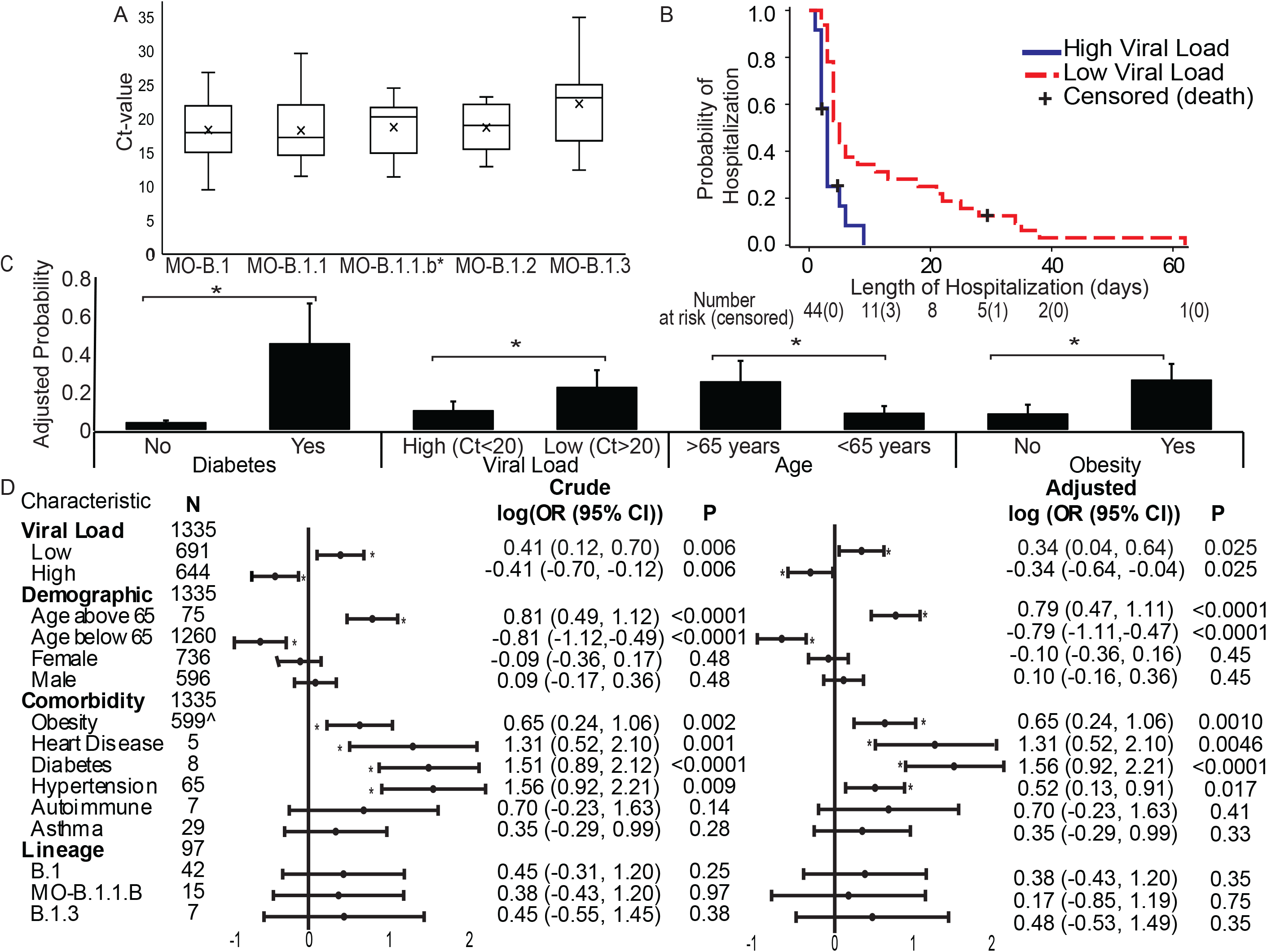
Clinical complications of genetic lineages and high viral loads in the nasopharyngeal swabs. A) Viral loads across Missouri lineages as measured by Ct on qRT-PCR assays. The boxes represent interquartile ranges, the horizontal lines within the boxes represent median Ct, and the whiskers denote the minimum and maximum values. All PANGOLIN-adapted Missouri lineages with at least five samples were included in the analysis; ^*^, novel mutations detected in Missouri. B) Kaplan-Meier curve for time from hospital admission to discharge in days for 44 hospitalized patients comparing high and low viral loads. C) A logistic regression was calculated to predict hospitalization based on patient demographics, comorbidities, and outcomes. The significant categorical predictors are reported here as adjusted probabilities denoted with standard error bars. A significant regression was found (p<0.0001), where age is categorized as above 65 or below 65 years, obesity coded as Yes=1, No=0, month of initial presentation in month number (ie. March = 3), diabetes coded as Yes=1, No=0, and viral load coded as High (Ct ≤20) or Low (Ct>20). Age, body mass index, month of initial presentation, diabetes, and viral load are significant predictors of hospitalization. ^*^, p<0.05. D) Forest plot of univariate logistic regressions for categorical factors tested for COVID-19 hospitalization. The panel on the right-hand side is adjusted for month of initial diagnosis. Log 10 of odds ratios (OR) and 95% confidence intervals (CI) are reported. ^^^, Obesity is out of 923 patients with available body mass index data. Intervals overlapping the value of 0 is considered not significant. ^*^, p<0.05.

### Increased viral load in clinical samples is correlated with elevated viral replication ability

Our observations of a temporal pattern to viral load and mutations in receptor binding domains of the S protein and RdRp complex led us to hypothesize that differences may have existed in virus replication properties. Eighteen isolates selected as representative sample strains from each viral load category and lineage were recovered from swab samples with Ct-values ranging from 9.09-37.76 (median=16.32) (Figure 3A). We determined growth kinetics for these isolates (Figure 3B) and observed a large diversity in viral growth patterns, especially during the initial 24 hours. Linear regression of viral proliferation (measured by Ct-values) at 24 hours showed that an increase of 1 cycle correlated with a decrease of 8.83 log_10_ (plaque forming units/mL) (Pearson correlation coefficient=-0.59, p=0.01) (Figure 3C). Taken together, the growth kinetics analyses revealed that strains with higher viral loads in clinical samples proliferated more efficiently than those with lower viral loads.

### High viral load is associated with decreased hospitalizations and shorter hospital lengths of stay

To understand potential clinical implications of the identified mutations, we analyzed associations among genetic variants in the Missouri lineages with disease severity (hospitalization and length of hospitalization) and viral loads in clinical samples. We found no significant associations between the Missouri viruses and viral loads (Chi-Square=3.35, p-value [p]=0.76) (Figure 4A) or hospitalizations (Chi-Square=0.33, p=1.00).

We next examined potential correlations between viral load in clinical swabs and outcomes. A univariable analysis was performed to test the effect of viral load, demographics, comorbidities, and lineages on the risk for hospitalization (Figure 4D). High viral load (Adjusted log odds ratio [ALOR]=-0.34, p=0.025), age >65 years (ALOR=0.79, p<0.0001), obesity (ALOR=0.65, p=0.001), heart disease (ALOR=1.31, p=0.005), diabetes (ALOR=1.56, p<0.0001), and hypertension (ALOR=0.52, p=0.02) were independently associated with hospitalization. Using a multivariable logistic regression model, we found that patients of older age (odds ratio [OR]=1.06, p<0.0001), earlier month of diagnosis (OR=0.69, p=0.0002), higher body mass index (BMI) (OR=1.037, p=0.04), diabetes (OR=0.06, p=0.001), and low viral load (OR=0.39, p=0.03) were more likely to be hospitalized (eTable 5, Figure 4C).

Importantly, patients with high viral loads at sampling had fewer hospitalizations (OR=0.39, p=0.01). Of hospitalized patients, those with high viral load were discharged sooner (hazard ratio = 2.9, p=0.03) (Figure 4B) compared to patients with low viral loads. There was no difference in time from symptom onset to COVID-19 test between low and high viral load groups (5.38±10.48 days and 4.25±4.43 days, respectively) or in time from symptom onset to hospital admission (10.03±10.48 days and 8.75±4.09 days, respectively).

## DISCUSSION

Studies assessing SARS-CoV-2 viral load as a marker for COVID-19 disease severity have been inconclusive. Early studies found that viral loads were correlated with age, disease stage, severity, progression, and mortality^32-37^. Most of these studies observed patients during the early stages of the pandemic and often investigated already hospitalized patients. Our study expanded these observations to thousands of patients over an eight-month period, March-October 2020, which was powered to detect that the average viral load increased over the study period. Furthermore, patients with high viral loads were less likely to become hospitalized than patients with low viral loads even after adjusting for month of diagnosis, age, obesity, and diabetes. Thus, although advances in treatment have improved patient outcomes, this was unlikely the only cause for reduced hospitalizations. Our results did suggest that heart disease and hypertension were confounding variables, not associated with hospitalization after accounting for other variables in the model. Additionally, patients with high viral loads were more likely to become discharged sooner than those with low viral loads. Because viral load in nasopharyngeal swabs typically declines after the first week of infection^38,39^, one confounding factor may have been delayed testing, especially at the beginning of the pandemic from limited access to testing. We did not, however, find differences in time from symptom onset to initial COVID-19 swab between the high and low viral load groups among hospitalized patients.

The clinical findings and demographics from our study are reflective of national data, lending confidence towards the generalizability of our patient population^1^. Exceptions include the distribution of cases within the 18-29-year age category where we noted 50% higher proportional incidence than CDC data; correspondingly, our >50 age categories were slightly less than national data. This reflects the catchment area of our study which includes the University of Missouri with a high proportion of college-aged students. Additionally, our population had slightly lower Asian, higher Black or African American and White, and over 50% lower Hispanic or Latino individuals than national data, reflecting the overall racial and ethnic distribution in Missouri. We also found that other risk factors for hospitalization included older age, elevated BMI, and diabetes mellitus, consistent with published studies^40,41^.

Genomic and phylogenetic evidence revealed multiple viral introductions into and across the state and numerous mutations during the first four months of the pandemic. We identified novel Missouri variants and lineages among both urban and rural communities. Four unique, well-supported sub-lineages were identified in Missouri, of which MO-B.1.1.b appeared to have persisted for at least two months. All four lineages emerged during the July sampling period in Southwestern Missouri and appeared more likely to arise in the urban Springfield area, then spread to neighboring urban and rural communities. Further studies are needed to evaluate whether any of these remained locally prominent.

Over the past year, variants containing the D614G and N501Y mutations dominated global outbreaks. D614G (B.1 lineage), first identified in January 2020, became predominant worldwide by June 2020^42^. The N501Y strain (B.1.1.7 lineage) was detected first in the United Kingdom^10^and then appeared in other countries^9^including the United States^11^. D614G mutations enhanced viral replication and transmission but not pathogenesis in laboratory settings^43,44^. However, disease transmission and severity of these novel variants in humans remain unclear^45^. Prior studies suggested that D614G is associated with higher viral load^14,46^, but the mutation was already predominant in both high and low viral load Missouri strains. In our findings, viral load increased over time (Figure 1). We explored whether a particular genetic variant was associated with the increased viral loads and were unable to find a clear association. Thus, we speculate that throughout the pandemic, all emerging variants of the virus were adapting to the human populations with greater viral replication efficiency. Of interest, multiple sites, especially at the Spike protein and RdRp complex, across multiple Missouri sub-lineages were under positive selection (Figure 3). Selection at the RdRp complex may affect viral replication and transcription^47,48^, while selection at the S protein may affect host receptor binding and viral entry^47^. Further examination of mutations from this and other studies will elucidate the phenotypic effects of these mutations.

Increasing case studies of reinfection and studies involving waning neutralizing antibody (Nab) titers raise concerns for herd immunity and long-lasting efficacy of vaccines^49-51^. CDC criteria for SARS-CoV-2 reinfection include persons with paired respiratory specimens at least 90 days apart and symptomatic persons 45-89 days after initial illness with respective respiratory specimens showing differing lineages^52^. Recent studies show Nab titers to SARS-CoV-2 decline as early as 23 days following initial infection^53^. In this study, a young female patient was identified with two genetically distinct SARS-CoV-2 strains within two weeks, indicating that re-infection can occur within a much shorter period than expected.

There are several limitations to this study. Analyses with viral loads are limited by variability in nasopharyngeal swabbing techniques, which may cause inconsistencies in Ct-values, although the same sampling and processing protocol was used throughout this study. Additionally, during the initial phases of the pandemic, testing was generally limited to patients with more severe symptoms, potentially skewing viral load findings. Despite these limitations, we analyzed a large, representative sample, and adjusted for these confounders.

In summary, multiple novel lineages were identified, and locally acquired mutations, present at both the urban and rural levels, remained predominant in the community. Although we were unable to find associations between specific variants and greater disease severity, Missouri COVID-positive individuals that presented with increased viral shedding had less severe disease by several measures. Continued monitoring of the impacts of these novel variants, particularly of those in the regions of vaccine targets, will be essential to the management of this pandemic.

## Supporting information

eMethods

## Data Availability

Genome data may be accessed using GenBank Accession Numbers: MW004168, MW521383-MW521516, MW525282. 

## Acknowledgments

Author contributions: XFW conceived this study, XFW, JAM, and CYT designed the analysis, CYT, YW, KS, TL, TH, and CS collected the data, DRS, RH, DR, CS, RT, GML, JH, and XFW contributed data or analysis tools, CYT, YW, CG, DRS, and XFW performed the analysis, CYT and XFW wrote the paper, RW, DRS, JAM, TH, CYT, and XFW revised the paper. XFW and CYT had full access to all the data in the study and takes responsibility for the integrity of the data and the accuracy of the data analysis.

The authors would like to thank Paige Beauparlant (UMHC), CoxHealth research coordinators, and Eren Ufuktepe and Katie Wilkinson from the UMHC Cerner Team for help with chart review, Simone Camp (UMHC) and Michelle Beckwith (PTC Labs) for help with sample collection and serving as the third-party honest brokers, and Varun Goel and Michael Emch (University of North Carolina) for feedback on geographical visualization methods. Additionally, we would like to thank the administrators and curators of the GISAID database, and research groups across the globe for supporting the rapid and transparent sharing of genomic data during the COVID-19 pandemic. The opinions expressed are the private views of the authors and are not to be conveyed as official or signifying the views of the Department of the Army or the Department of Defense. Authors have no conflicts of interest to report.

## Data Sharing Statement

Genome data may be accessed using GenBank Accession Numbers: MW004168, MW521383-MW521516, MW525282.

## References

1. CDC COVID Data Tracker. Updated 01-28-2021. https://covid.cdc.gov/covid-data-tracker/ (accessed 01-28-21).

2. Woolf SH, Chapman DA, Lee JH. COVID-19 as the Leading Cause of Death in the United States. JAMA 2021;325(2):123–124. doi:10.1001/jama.2020.24865.

3. Zhao WM, Song SH, Chen ML, et al. The 2019 novel coronavirus resource. PMID: 32102777 DOI: 10.16288/j.yczz.20-030.

4. Koyama T, Platt D, Parida L. Variant analysis of SARS-CoV-2 genomes. Bulletin of the World Health Organization 2020; 98(7): 495–504.

5. van Dorp L, Acman M, Richard D, et al. Emergence of genomic diversity and recurrent mutations in SARS-CoV-2. Infection, Genetics and Evolution 2020; 83: 104351.

6. Rambaut A, Holmes EC, O’Toole Á, et al. A dynamic nomenclature proposal for SARS-CoV-2 lineages to assist genomic epidemiology. Nature Microbiology 2020; 5(11): 1403–7.

7. Velazquez-Salinas L, Zarate S, Eberl S, Gladue DP, Novella I, Borca MV. Positive Selection of ORF1ab, ORF3a, and ORF8 Genes Drives the Early Evolutionary Trends of SARS-CoV-2 During the 2020 COVID-19 Pandemic. Frontiers in Microbiology 2020; 11: 550674.

8. Cagliani RA-O, Forni DA-O, Clerici M, Sironi M. Computational Inference of Selection Underlying the Evolution of the Novel Coronavirus, Severe Acute Respiratory Syndrome Coronavirus 2. Journal of Virology 2020; 10.1128/JVI.00411-20. (1098–5514 (Electronic)).

9. Tegally H, Wilkinson E, Giovanetti M, et al. Emergence and rapid spread of a new severe acute respiratory syndrome-related coronavirus 2 (SARS-CoV-2) lineage with multiple spike mutations in South Africa. medRxiv 2020: 2020.12.21.20248640.

10. Leung K, Shum MH, Leung GM, Lam TT, Wu JT. Early empirical assessment of the N501Y mutant strains of SARS-CoV-2 in the United Kingdom, October to November 2020. medRxiv 2020: 2020.12.20.20248581.

11. Tu H, Avenarius MR, Kubatko L, et al. Distinct Patterns of Emergence of SARS-CoV-2 Spike Variants including N501Y in Clinical Samples in Columbus Ohio. bioRxiv 2021: 10.1101/2021.01.12.426407.

12. Plante JA, Liu Y, Liu J, et al. Spike mutation D614G alters SARS-CoV-2 fitness. Nature 2020. https://doi.org/10.1038/s41586-020-2895-3.

13. Yurkovetskiy L, Wang X, Pascal KE, et al. Structural and Functional Analysis of the D614G SARS-CoV-2 Spike Protein Variant. Cell 2020; 183(3): 739-51.e8.

14. Korber B, Fischer WM, Gnanakaran S, et al. Tracking Changes in SARS-CoV-2 Spike: Evidence that D614G Increases Infectivity of the COVID-19 Virus. Cell 2020; 10.1016/j.cell.2020.06.043. (1097–4172 (Electronic)).

15. ShowMeStrong Recovery Plan: Public Health Dashboard. State of Missouri. Updated 12/26/2020. https://showmestrong.mo.gov/data/public-health/ (accessed 12/27/2020).

16. Lu X Fau - Wang L, Wang L Fau - Sakthivel SK, Sakthivel Sk Fau - Whitaker B, et al. US CDC Real-Time Reverse Transcription PCR Panel for Detection of Severe Acute Respiratory Syndrome Coronavirus 2. Emerging Infectious Diseases 2020; 10.3201/eid2608.201246 (1080–6059 (Electronic)).

17. Li T, Chung HK, Pireku PK, et al. Rapid High Throughput Whole Genome Sequencing of SARS-CoV-2 by using One-step RT-PCR Amplification with Integrated Microfluidic System and Next-Gen Sequencing. bioRxiv 2020: 10.1101/2020.11.04.369165.

18. Shu Y, McCauley J. GISAID: Global initiative on sharing all influenza data - from vision to reality. Eurosurveillance 2017; 10.2807/1560-7917.ES.2017.22.13.30494.

19. Wan XF, Chen G, Luo F, Emch M, Donis R. A quantitative genotype algorithm reflecting H5N1 Avian influenza niches. Bioinformatics 2007; 23(18): 2368–75.

20. Wan XF, Ozden M, Lin G. Ubiquitous reassortments in influenza A viruses. Journal of Bioinformatics and Computional Biology 2008; 6(5): 981–99.

21. Zhao ZM, Shortridge KF, Garcia M, Guan Y, Wan XF. Genotypic diversity of H5N1 highly pathogenic avian influenza viruses. Journal of General Virology 2008; 89(Pt 9): 2182–93.

22. Wu X, Wan XF, Wu G, Xu D, Lin G. Phylogenetic analysis using complete signature information of whole genomes and clustered Neighbour-Joining method. International Journal of Bioinformatics Research and Applications 2006; 2(3): 219–48.

23. Wu X, Goebel R, Wan XF, Lin G. Whole genome composition distance for HIV-1 genotyping. Computational Systems Bioinformatics Conference 2006: 179–90.

24. Lin G, Cai Z, Wu J, Wan XF, Xu L, Goebel R. Identifying a few foot-and-mouth disease virus signature nucleotide strings for computational genotyping. BMC Bioinformatics 2008; 9: 279.

25. Edgar RC. MUSCLE: multiple sequence alignment with high accuracy and high throughput. Nucleic Acids Research 2004; 10.1093/nar/gkh340. (1362-4962 (Electronic)).

26. Bouckaert R, Heled J, Kühnert D, et al. BEAST 2: a software platform for Bayesian evolutionary analysis. PLoS Computational Biology 2014; 10(4): e1003537.

27. Bielejec F, Baele G, Vrancken B, Suchard MA, Rambaut A, Lemey P. SpreaD3: Interactive Visualization of Spatiotemporal History and Trait Evolutionary Processes. Molecular Biology and Evolution 2016; 33(8): 2167–9.

28. Yang Z. PAML 4: Phylogenetic Analysis by Maximum Likelihood. Molecular Biology and Evolution 2007; 24(8): 1586–91.

29. Lole KS, Bollinger RC, Paranjape RS, et al. Full-length human immunodeficiency virus type 1 genomes from subtype C-infected seroconverters in India, with evidence of intersubtype recombination. Journal of Virology 1999; 73(1): 152–60.

30. ShowMeStrong Recovery Plan: Data Downloads. State of Missouri. Updated 11/27/2020. https://showmestrong.mo.gov/data-download/ (accessed 11/27/2020).

31. A Review of the SARS-CoV-2 (COVID-19) Genome and Proteome. April 21, 2020. https://www.genetex.com/MarketingMaterial/Index/SARS-CoV-2_Genome_and_Proteome (accessed February 1, 2021).

32. To KK, Tsang OT, Leung WS, et al. Temporal profiles of viral load in posterior oropharyngeal saliva samples and serum antibody responses during infection by SARS-CoV-2: an observational cohort study. Lancet Infectious Diseases 2020; 20(5): 565–74.

33. Zheng S, Fan J, Yu F, et al. Viral load dynamics and disease severity in patients infected with SARS-CoV-2 in Zhejiang province, China, January-March 2020: retrospective cohort study. BMJ 2020; 369: m1443.

34. Yu X, Sun S, Shi Y, Wang H, Zhao R, Sheng J. SARS-CoV-2 viral load in sputum correlates with risk of COVID-19 progression. Critical Care 2020; 24(1): 170.

35. Liu Y, Yan LM, Wan L, et al. Viral dynamics in mild and severe cases of COVID-19. Lancet Infectious Diseases 2020; 20(6): 656–7.

36. Guallar MP, Meiriño R, Donat-Vargas C, Corral O, Jouvé N, Soriano V. Inoculum at the time of SARS-CoV-2 exposure and risk of disease severity. International Journal of Infectious Diseases 2020; 97: 290–2.

37. Pujadas E, Chaudhry F, McBride R, et al. SARS-CoV-2 viral load predicts COVID-19 mortality. The Lancet Respiratory Medicine 2020; 8(9): e70.

38. Shi F, Wu T, Zhu X, et al. Association of viral load with serum biomakers among COVID-19 cases. Virology 2020; 546: 122–6.

39. Kampf G, Brüggemann Y, Kaba HEJ, et al. Potential sources, modes of transmission and effectiveness of prevention measures against SARS-CoV-2. Journal of Hospital Infection 2020; 106(4): 678–97.

40. Ortiz-Prado E, Simbaña-Rivera K, Gómez-Barreno L, et al. Clinical, molecular, and epidemiological characterization of the SARS-CoV-2 virus and the Coronavirus Disease 2019 (COVID-19), a comprehensive literature review. Diagnostic Microbioly and Infectious Disease 2020; 98(1): 115094.

41. Li X, Xu S, Yu M, et al. Risk factors for severity and mortality in adult COVID-19 inpatients in Wuhan. Journal of Allergy and Clinical Immunology 2020; 146(1): 110–8.

42. SARS-CoV-2 Variants. World Health Organizaiton. Updated 12/31/2020. https://www.who.int/csr/don/31-december-2020-sars-cov2-variants/en/ (accessed 1/17/2021).

43. Zhang L, Jackson CB, Mou H, et al. SARS-CoV-2 spike-protein D614G mutation increases virion spike density and infectivity. Nature Communications 2020; 11(1): 6013.

44. Hou YJ, Chiba S, Halfmann P, et al. SARS-CoV-2 D614G variant exhibits efficient replication ex vivo and transmission in vivo. Science 2020; 370(6523): 1464.

45. Volz E, Hill V, McCrone JT, et al. Evaluating the Effects of SARS-CoV-2 Spike Mutation D614G on Transmissibility and Pathogenicity. Cell 2021; 184(1): 64-75.e11.

46. Lorenzo-Redondo R, Nam HH, Roberts SC, et al. A clade of SARS-CoV-2 viruses associated with lower viral loads in patient upper airways. EBioMedicine 2020; 10.1016/j.ebiom.2020.103112 PMCID: PMC7655495. (2352–3964 (Electronic)).

47. Dhama K, Khan S, Tiwari R, et al. Coronavirus Disease 2019-COVID-19. Clinical Microbioly Reviews 2020; 33(4).

48. Jiang Y, Yin W, Xu HE. RNA-dependent RNA polymerase: Structure, mechanism, and drug discovery for COVID-19. Biochemical Biophysical Research Communications 2020: S0006-291X(20)31721-6.

49. Cohen JI, Burbelo PD. Reinfection with SARS-CoV-2: Implications for Vaccines. Clinical Infectious Diseases 2020; 10.1093/cid/ciaa1866.

50. Muecksch F, Wise H, Batchelor B, et al. Longitudinal analysis of serology and neutralizing antibody levels in COVID19 convalescents. Journal of Infectious Diseases 2020; 10.1093/infdis/jiaa659. (1537–6613 (Electronic)).

51. Crawford KHD, Dingens AS, Eguia R, et al. Dynamics of neutralizing antibody titers in the months after SARS-CoV-2 infection. Journal of Infectious Diseases 2020; 10.1093/infdis/jiaa618. (1537–6613 (Electronic)).

52. Investigative Criteria for Suspected Cases of SARS-CoV-2 Reinfection (ICR). Center for Disease Control and Prevention. https://www.cdc.gov/coronavirus/2019-ncov/php/invest-criteria.html. Updated 10/27/2020 (accessed 12/27/20).

53. Seow JA-O, Graham C, Merrick BA-O, et al. Longitudinal observation and decline of neutralizing antibody responses in the three months following SARS-CoV-2 infection in humans. Nature Microbiology 2020; 10.1038/s41564-020-00813-8. (2058–5276 (Electronic)).

