## Supplementary material for "Increased SAR-CoV-2 shedding associated with reduced disease severity despite continually emerging genetic variants": eMethods

**Supplementary Information**

**Continually emerging SARS-CoV-2 genetic variants during COVID-19 pandemic increased viral shedding but caused less severe disease**

Cynthia Y. Tang^1,2,3^, Yang Wang^1,3^, Cheng Gao^1,3,4^, David R. Smith^5^, Jane A. McElroy^1^, Tao Li^6^, Karen Segovia^1,3^, Tricia Haynes^1^, Richard Hammer^1^, Christopher Sampson^1^, Detlef Ritter^1^, Christopher Schulze^7^, Robin Trotman^7^, Grace M Lidl^6^, Richard Webby^8^, Jun Hang^6^, Xiu-Feng Wan^1,2,3,4*^

^1^School of Medicine, University of Missouri, Columbia, MO, USA**;** ^2^Institute for Data Science and Informatics, University of Missouri, Columbia, MO, USA; ^3^Bond Life Sciences Center, University of Missouri, Columbia, MO, USA; ^4^College of Engineering, University of Missouri, Columbia, MO, USA; ^5^College of Veterinary Medicine, Mississippi State University, Mississippi State, MS, USA; ^6^Walter Reed Army Institute of Research, Silver Spring, MD, USA; ^7^CoxHealth Center, Springfield, MO, USA; ^8^Department of Infectious Diseases, St. Jude Children's Research Hospital, Memphis, TN, USA.

**eMETHODS**

**Chart review:** Comorbidities included heart failure, heart disease, hypertension, atrial fibrillation or flutter, diabetes mellitus, asthma, cancer, chronic kidney disease, acute kidney injury, human immunodeficiency virus/acquired immunodeficiency syndrome (HIV/AIDs), mental health disorders, coagulation or blood disorder, autoimmune disease, history of transplant, obesity, pregnancy, or peripheral vascular disease. Demographics included age, race, sex, and zip code, disease course information included date of initial diagnosis, symptoms, and time from symptom onset to diagnosis and hospitalization, and hospital information included status of hospitalization (yes/no) and length of stay if hospitalized.

**Viral isolation and growth kinetics**: For virus isolation, 10-fold diluted clinical samples were passaged on Vero E6 (CRL1586™, ATCC) or Vero (NR-10385, BEI Resources) cells a maximum of three times until cytopathic effect was observed. To determine virus replication kinetics, Vero E6 cells were seeded on 6-well plates (USA Scientific). The next day, cells at a confluence of ~90% were inoculated with a testing virus isolate at a multiplicity of infection (MOI) of 0.01. After one hour of adsorption, the inoculum was removed, and the cells were washed with 1xDulbecco's Phosphate Buffered Saline (DPBS) and covered with fresh optiMEM (Gibco, Thermo Fisher Scientific). The supernatants were harvested at 12, 24, 48, and 72 hours post inoculation and then titrated by using plaque assays. For the plaque assays, 10-fold serially diluted viruses were added onto Vero E6 cells and incubated for one hour. After removal of the inoculums, followed by washes with 1xDPBS, the cells were overlaid with a 1:1 mixture of 1.2 % low melting point agarose (Lonza) and 2xMinimum Essential Media (Gibco, Thermo Fisher Scientific) containing bovine serum albumin (Sigma‐Aldrich) with a final concentration of 0.3% and incubated for three days. Thereafter, the cells were fixed and stained with 0.03% neutral red (Sigma‐Aldrich). The plaques on the monolayer were then counted to calculate the viral titer.

**Genetic sequencing and assembly**. SARS-CoV-2 whole genome RT-PCR amplification was conducted either using the Access Array (AA) system (Fluidigm Corporation, CA, USA) with 35 pairs of customer designed specific primers targeting the reference sequence NC_045512.2, or using the two step RT-PCR ARTIC protocol version 3 (24-Mar-2020)^1^ with v3 primers pool 1 and pool 2 (https://artic.network/ncov-2019). Amplicon libraries were then prepared using the Nextera DNA Flex Library Prep kit, followed by sequencing with MiSeq Reagent Kit v3 (600-cycle) and MiSeq sequencing system (Illumina, San Diego, CA, USA).

The quality of paired-end reads obtained from MiSeq sequencing were analyzed and consensus sequences were constructed using Qiagen CLC Genomics Workbench 20.0.4 and the Qiagen “Analysis of SARS-CoV-2 using MinION sequences” protocol was used in genetic variant analyses. Sequences were imported as paired reads and trimmed with a quality score of 0.05. The trimmed reads were mapped to the reference genome, NC_04412.2. A minimum coverage of 10 was required to assemble the consensus sequences. Non-synonymous amino acid variants were identified with a variant probability threshold of 80% and at least 2 counts. Sequences with less than 80% coverage were excluded from analysis. Mutations were confirmed manually using BioEdit v7.2.5.

**Phylogenetic and phylogeographic analyses.** To identify the potential precursor viruses for Missouri strains, all of the 110,901 SARS-CoV-2 complete genomes available on GISAID as of September 20, 2020 were downloaded. A total of 297 of the genetically closest genomic sequences to Missouri samples were selected using alignment-free complete composition vector^2-7^ to further examine evolutionary relations among these viruses. Sequences were aligned using MUSCLE^8^, and phylogenetic analyses with BEAST2^9^ with a Hasegawa-Kishino-Yano (HKY) substitution model (k=2.0) with empirical frequencies, strict clock model, and Coalescent Constant Population prior was used. Markov chain Mote Carlo (MCMC) was used with a chain length of 500,000,000 stored every 50000 and pre burn-in of 10%. The results were analyzed in Tracer v1.7.1 and convergence was assessed with a cutoff of 200 for the effective sample size (ESS). The consensus tree was generated using TreeAnnotator v2.6.3.0. The trees were visualized with FigTree v1.4.4 (<http://tree.bio.ed.ac.uk/software/figtree/>). All posterior probabilities of > 70% and PANGOLIN (Phylogenetic Assignment of Named Global Outbreak LINeages) lineages with greater five sequences were annotated. Lineages were additionally supported by overall mean distances and distance matrices between sequences, which were calculated using Molecular Evolutionary Genetics Analysis (MEGA X) v.10.1.8^10^ with a bootstrap of 1,000.

**Positive selection analyses.** The aligned sequences from the phylogeny were used to analyze SARS-CoV-2 evolution at the following proteins: ORF1ab (including nsp 1-16 except for nsp 11 as it is fully encompassed by nsp 12), S, ORF3a, E, M, ORF8, N, ORF10. Only complete sequences at each region were included in the analysis. Amino acid files were created using BioEdit v7.2.5 and newick tree files were constructed using MEGA v10.1.8 using an HKY substitution model and bootstrap of 1,000. CodeML of Phylogenetic Analysis by the Maximum Likelihood (PAML) v4.9 software was used for analysis^11^. A dN/dS (nonsynonymous to synonymous mutations) of 1 indicates neutral evolution, > 1 indicates positive selection, and < 1 indicates negative selection. Sequence conservation was visualized with SimPlot^12^

**SUPPLEMENTARY TABLES**

**eTable 1. Sequences submitted to GenBank**

| **Sample ID** | **GeneBank Accession ID** | **Sample ID** | **GeneBank Accession ID** |
| --- | --- | --- | --- |
| 1425xA10 | MW521383 | 1508xG11 | MW521450 |
| 1425xE1 | MW521384 | 1514xE7 | MW521451 |
| 1432xE3 | MW521385 | 1515xH11 | MW521452 |
| 1432xH8 | MW521386 | 1515xH4 | MW521453 |
| 1436xB10 | MW521387 | 1516xB10 | MW521454 |
| 1436xE10 | MW521388 | 1516xC4 | MW521455 |
| 1436xE11 | MW521389 | 1516xD7 | MW521456 |
| 1436xE6 | MW521390 | 1516xF10 | MW521457 |
| 1436xE8 | MW521391 | 1516xG4 | MW521458 |
| 1436xH6 | MW521392 | 1516xG7 | MW521459 |
| 1437xE9 | MW521393 | 1518xB3 | MW521460 |
| 1437xH6 | MW521394 | 1518xE12 | MW521461 |
| 1438xA11 | MW521395 | 1518xG10 | MW521462 |
| 1438xA5 | MW521396 | 1529xC1 | MW521463 |
| 1452xG12 | MW521397 | 1529xF6 | MW521464 |
| 1452xG2 | MW521398 | 1529xF8 | MW521465 |
| 1452xH9 | MW521399 | 1530xF7 | MW521466 |
| 1453xB8 | MW521400 | 1532xE1 | MW521467 |
| 1453xB9 | MW521401 | 1573xB2 | MW521468 |
| 1453xE2 | MW521402 | 20x1001 | MW521469 |
| 1453xF4 | MW521403 | 20x1003 | MW521470 |
| 1453xH10 | MW521404 | 20x1005 | MW521471 |
| 1453xH7 | MW521405 | 20x1007 | MW521472 |
| 1463xC12 | MW521406 | 20x1012 | MW521473 |
| 1463xC2 | MW521407 | 20x1014 | MW521474 |
| 1463xD12 | MW521408 | 20x1016 | MW521475 |
| 1466xB11 | MW521409 | 20x1019 | MW521476 |
| 1466xB3 | MW521410 | 20x1020 | MW521477 |
| 1466xD5 | MW521411 | 20x1023 | MW521478 |
| 1466xF2 | MW521412 | 20x1025 | MW521479 |
| 1466xF3 | MW521413 | 20x1029 | MW521480 |
| 1466xH2 | MW521414 | 20x1030 | MW521481 |
| 1469xA1 | MW521415 | 20x1031 | MW521482 |
| 1475xC11 | MW521416 | 20x1032 | MW521483 |
| 1475xC9 | MW521417 | 20x1036 | MW521484 |
| 1475xD8 | MW521418 | 20x1039 | MW521485 |
| 1475xE3 | MW521419 | 20x1043 | MW521486 |
| 1479xG9 | MW521420 | 20x1047 | MW521487 |
| 1480xH12 | MW521421 | 20x1049 | MW521488 |
| 1481xA11 | MW521422 | 20x1050 | MW521489 |
| 1481xD2 | MW521423 | 20x1052 | MW521490 |
| 1481xH5 | MW521424 | 20x1057 | MW521491 |
| 1482xB4 | MW521425 | 20x1062 | MW521492 |
| 1482xE4 | MW521426 | 20x1063 | MW521493 |
| 1483xA7 | MW521427 | 20x1067 | MW521494 |
| 1483xF2 | MW521428 | 20x1068 | MW521495 |
| 1484xA3 | MW521429 | 20x1084 | MW521496 |
| 1484xB12 | MW521430 | 20x1085 | MW521497 |
| 1484xE11 | MW521431 | 20x1088 | MW521498 |
| 1484xE9 | MW521432 | 20x1089 | MW521499 |
| 1499xA4 | MW521433 | 20x1094 | MW521500 |
| 1499xC10 | MW521434 | 20x1099 | MW521501 |
| 1499xC11 | MW521435 | 20x1104 | MW521502 |
| 1499xD3 | MW521436 | 20x1105 | MW521503 |
| 1499xF5 | MW521437 | 20x1106 | MW521504 |
| 1499xF7 | MW521438 | 20x1109 | MW521505 |
| 1499xH8 | MW521439 | 20x1118 | MW521506 |
| 1500xF5 | MW521440 | 1445xD9 | MW521507 |
| 1501xA6 | MW521441 | 20x1035 | MW521508 |
| 1501xF7 | MW521442 | 20x1064 | MW521509 |
| 1501xG3 | MW521443 | 20x1009 | MW521510 |
| 1502xA3 | MW521444 | 20x1011 | MW521511 |
| 1502xB4 | MW521445 | 20x1048 | MW521512 |
| 1502xD12 | MW521446 | 20x1051 | MW521513 |
| 1502xE7 | MW521447 | 20x1065 | MW521514 |
| 1503xA7 | MW521448 | 20x1096 | MW521515 |
| 1508xD10 | MW521449 | 20x1108 | MW521516 |
| MO-MU-1087 | MW004168 | 1446xH9 | MW525282 |

**eTable 2. Clade percent identities.**

| **Clade** | **Mean distance (d)** | **Standard Error** | **% Identity** |
| --- | --- | --- | --- |
| B | 0.0001825619 | 0.0000415155 | 99.982% |
| B.6 | 0.0001377372 | 0.0000344925 | 99.986% |
| A.3 | 0.0002532381 | 0.0000416374 | 99.975% |
| A | 0.0003537849 | 0.0000481421 | 99.965% |
| A.1 | 0.0002829375 | 0.0000394942 | 99.972% |
| B.1.1 | 0.0002986042 | 0.0000345816 | 99.970% |
| MO-B.1.1.a | 0.0000756191 | 0.0000241967 | 99.992% |
| MO-B.1.1.b | 0.0000851617 | 0.0000272789 | 99.991% |
| B.1.1.10 | 0.0000905780 | 0.0000235853 | 99.991% |
| B.1 | 0.0003065711 | 0.0000271180 | 99.969% |
| B.1.5 | 0.0001906879 | 0.0000408516 | 99.981% |
| MO-B.1.c | 0.0002373017 | 0.0000583934 | 99.976% |
| B.1.3 | 0.0002928698 | 0.0000494315 | 99.971% |
| B.1.2 | 0.0004423176 | 0.0000479856 | 99.956% |
| MO-B.1.2.d | 0.0000873197 | 0.0000321116 | 99.991% |
| Full Tree | 0.0004978965 | 0.0000446443 | 99.950% |

Sequence identities of each clade. Overall mean p-distance (d) and standard error was calculated using MEGA X. Percent identity was calculated with (1-d)*100.

**eTable 3. New nonsynonymous mutations in Missouri samples.**

| **Mutation** | **Frequency** |
| --- | --- |
| NSP1 | |
| R24C | 2 |
| L92F | 1 |
| NSP2 | |
| R107S | 3 |
| D155N | 2 |
| L274F | 1 |
| D286N | 1 |
| T332I | 1 |
| G344V | 2 |
| K384N | 1 |
| V426F | 1 |
| K493K | 1 |
| C509G | 1 |
| S512F | 1 |
| NSP3 | |
| P74S | 2 |
| F90S | 1 |
| E122D | 1 |
| D135Y | 2 |
| M196I | 1 |
| H342Q | 5 |
| A480V | 1 |
| A602V | 1 |
| L620F | 1 |
| V747L | 1 |
| Q764H | 1 |
| T808I | 1 |
| P822S | 5 |
| P874S | 1 |
| A876V | 1 |
| P992L | 1 |
| P1103S | 1 |
| N1178T | 6 |
| A1179T | 6 |
| S1212P | 1 |
| M1218I | 1 |
| P1260S | 2 |
| D1318N | 1 |
| P1326L | 1 |
| M11441I | 1 |
| K1679N | 2 |
| I1751T | 1 |
| M1788I | 2 |
| NSP4 | |
| M33I | 2 |
| T37I | 2 |
| T189I | 1 |
| T214K | 1 |
| A230V | 1 |
| A286V | 1 |
| M366I | 18 |
| A446V | 1 |
| S481L | 1 |
| T492I | 1 |
| NSP5 | |
| K89R | 2 |
| T135I | 1 |
| A191V | 1 |
| NSP6 | |
| L19F | 1 |
| L142I | 1 |
| V178I | 1 |
| NSP7 | |
| M75I | 1 |
| NSP8 | |
| K36Q* | 1 |
| T145I* | 1 |
| NSP10 | |
| M137V | 1 |
| NSP12 | |
| A2V | 11 |
| C22F | 18 |
| D92Y | 2 |
| A423V | 1 |
| T591I | 1 |
| D738Y | 2 |
| NSP13 | |
| P77L | 1 |
| T172I* | 1 |
| E341D | 1 |
| T431I* | 2 |
| K460R* | 1 |
| S468L* | 3 |
| NSP14 | |
| T16I | 1 |
| T113I | 1 |
| A281V | 2 |
| P297L | 2 |
| K311N | 1 |
| V381L | 1 |
| NSP15 | |
| Q196H | 1 |
| S241I | 1 |
| P262L | 6 |
| NSP16 | |
| L212F | 3 |
| I267T | 1 |
| S | |
| H49Y | 1 |
| S98F | 1 |
| Y248H | 1 |
| W258G | 1 |
| T306I | 1 |
| S477N | 1 |
| S591F | 1 |
| A653V | 3 |
| L938F | 1 |
| M1050I | 1 |
| K1073N | 1 |
| D1153Y | 1 |
| D1163Y | 1 |
| ORF3a | |
| V50I | 8 |
| A99S | 1 |
| G100C | 1 |
| G174V | 2 |
| V225F | 1 |
| T229I | 1 |
| E | |
| V5G | 1 |
| F20S | 1 |
| P71S | 1 |
| L73F | 7 |
| M | |
| A2V | 1 |
| A68D | 1 |
| G78C | 1 |
| T208I | 1 |
| D209Y | 1 |
| ORF8 | |
| K44E | 1 |
| S47F | 12 |
| N | |
| Q9H | 1 |
| D128Y | 1 |
| T135I | 1 |
| S190I | 3 |
| S232R | 1 |
| V270L | 10 |
| T334I | 1 |
| P364L | 1 |
| K373N | 1 |
| T391I | 1 |
| A419S | 1 |
| ORF10 | |
| I4L | 1 |

Novel Missouri mutations compared with precursor viruses included in this study. *, sites undergoing positive selection.

**eTable 4. Positively selected sites on each gene calculated using CodeML.**

| **Protein** | **Position** | **Original NC_045512.2** | **Pr(w>1)** | **dS/dN** |
| --- | --- | --- | --- | --- |
| **ORF1ab** | | | | |
| NSP8 | 27 | A | 1.000** | 4.756 |
|  | 36 | K | 1.000** | 4.756 |
|  | 55 | M | 1.000** | 4.756 |
|  | 79 | K | 1.000** | 4.756 |
|  | 129 | M | 1.000** | 4.756 |
|  | 145 | T | 1.000** | 4.756 |
| NSP12 | 314 | P | 0.998** | 24.286 |
| NSP13 | 47 | P | 1.000** | 2.137 |
|  | 73 | K | 1.000** | 2.137 |
|  | 127 | T | 1.000** | 2.137 |
|  | 153 | T | 1.000** | 2.137 |
|  | 169 | V | 1.000** | 2.137 |
|  | 206 | G | 1.000** | 2.137 |
|  | 224 | Y | 1.000** | 2.137 |
|  | 244 | E | 1.000** | 2.137 |
|  | 431 | T | 1.000** | 2.137 |
|  | 460 | K | 1.000** | 2.137 |
|  | 468 | S | 1.000** | 2.137 |
|  | 475 | F | 1.000** | 2.137 |
|  | 481 | T | 1.000** | 2.137 |
|  | 485 | S | 1.000** | 2.137 |
|  | 504 | P | 1.000** | 2.137 |
|  | 541 | Y | 1.000** | 2.137 |
| **S** | | | | |
| S | 682 | R | 0.998** | 18.323 |
|  | 1163 | D | 0.954* | 17.56 |

*, Probability > 95%; **, Probability > 99%.

**eTable 5. Logistic regression of COVID-19 hospitalizations.**

| **Regression Model** | | | | | |
| --- | --- | --- | --- | --- | --- |
|  | **DF** | **Estimate** | **Standard Error** | **Wald Chi-Square** | **Pr > ChiSq** |
| **Intercept** | 1 | -1.1484 | 1.3282 | 0.7476 | 0.3872 |
| **Age** | 1 | 0.056 | 0.00954 | 34.4367 | <.0001 |
| **Month** | 1 | -0.3719 | 0.1 | 13.8204 | 0.0002 |
| **Diabetes** | 1 | -2.8279 | 0.8778 | 10.3787 | 0.0013 |
| **BMI** | 1 | 0.0361 | 0.0179 | 4.0574 | 0.044 |
| **Viral Load (high)** | 1 | -0.9342 | 0.378 | 6.108 | 0.0135 |

| **Odds Ratio Estimates** | | | |
| --- | --- | --- | --- |
| **Effect** | **Point Estimate** | **95% Wald** | |
|  |  | **Confidence Limits** | |
| **Age** | 1.058 | 1.04 | 1.078 |
| **Month** | 0.689 | 0.57 | 0.839 |
| **Diabetes** | 0.059 | 0.01 | 0.33 |
| **BMI** | 1.037 | 1.00 | 1.074 |
| **Viral Load (high)** | 0.393 | 0.19 | 0.824 |

**SUPPLEMENTARY FIGURES**

**eFigure 1. Spatiotemporal spread of viral load for individual** **COVID-19 cases across Southwest and Central Missouri (March 17, 2020-October 31, 2020)**.

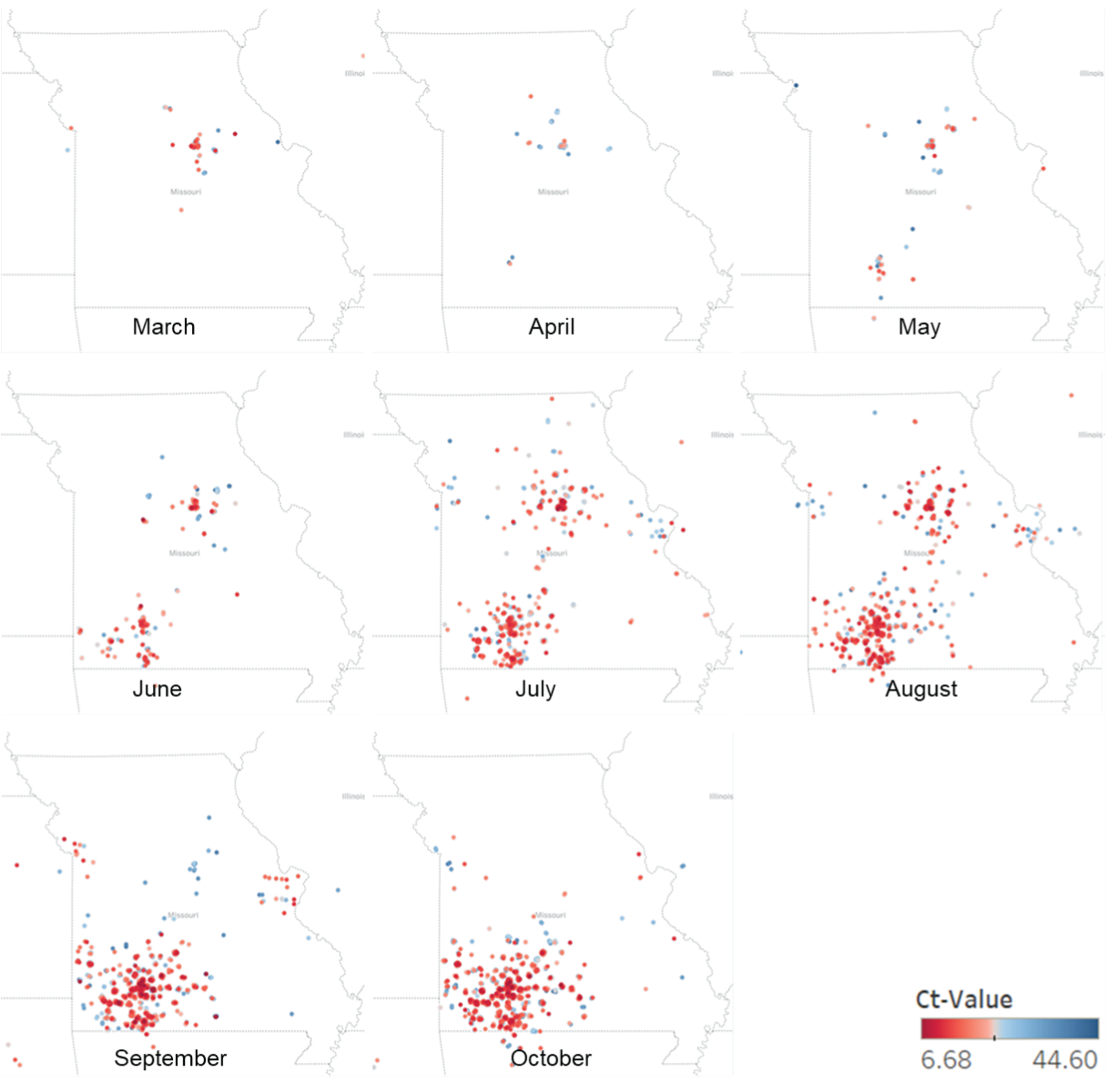

Each dot represents a new case. Red indicates higher viral load and blue indicates lower viral load detected in nasopharyngeal swabs by quantitative RT-PCR (qRT-PCR).

**eFigure 2. Phylogeny of SARS-CoV-2 viruses using whole genomic sequences.**

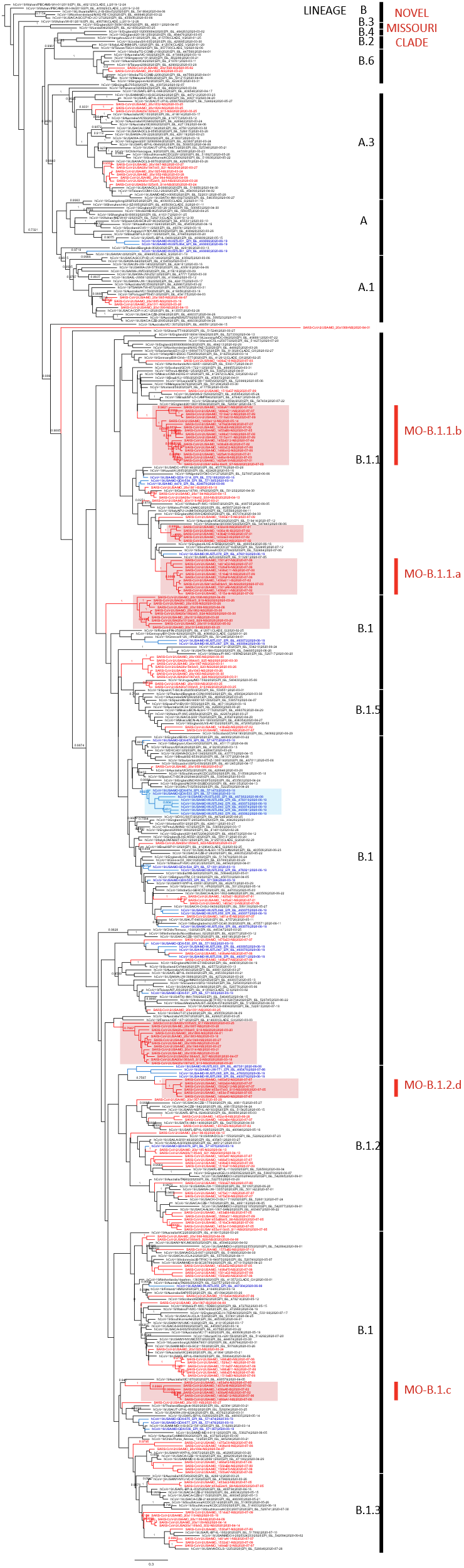

Bayesian tree of all patient nasopharyngeal swab samples and isolates (red), other Missouri samples previously uploaded to GISAID (blue), and other closely related sequences from GISAID (black) rooted to hCoV-19/Wuhan/IPBCAMS-WH-01/2019 (EPI ISL 402123). Annotated with taxa names and posterior probabilities > 0.70. Effective Sample Size (ESS) = 720.

**eFigure 3. Phylogeographic analysis of emergence and dispersion of SARS-CoV-2 in Missouri**.

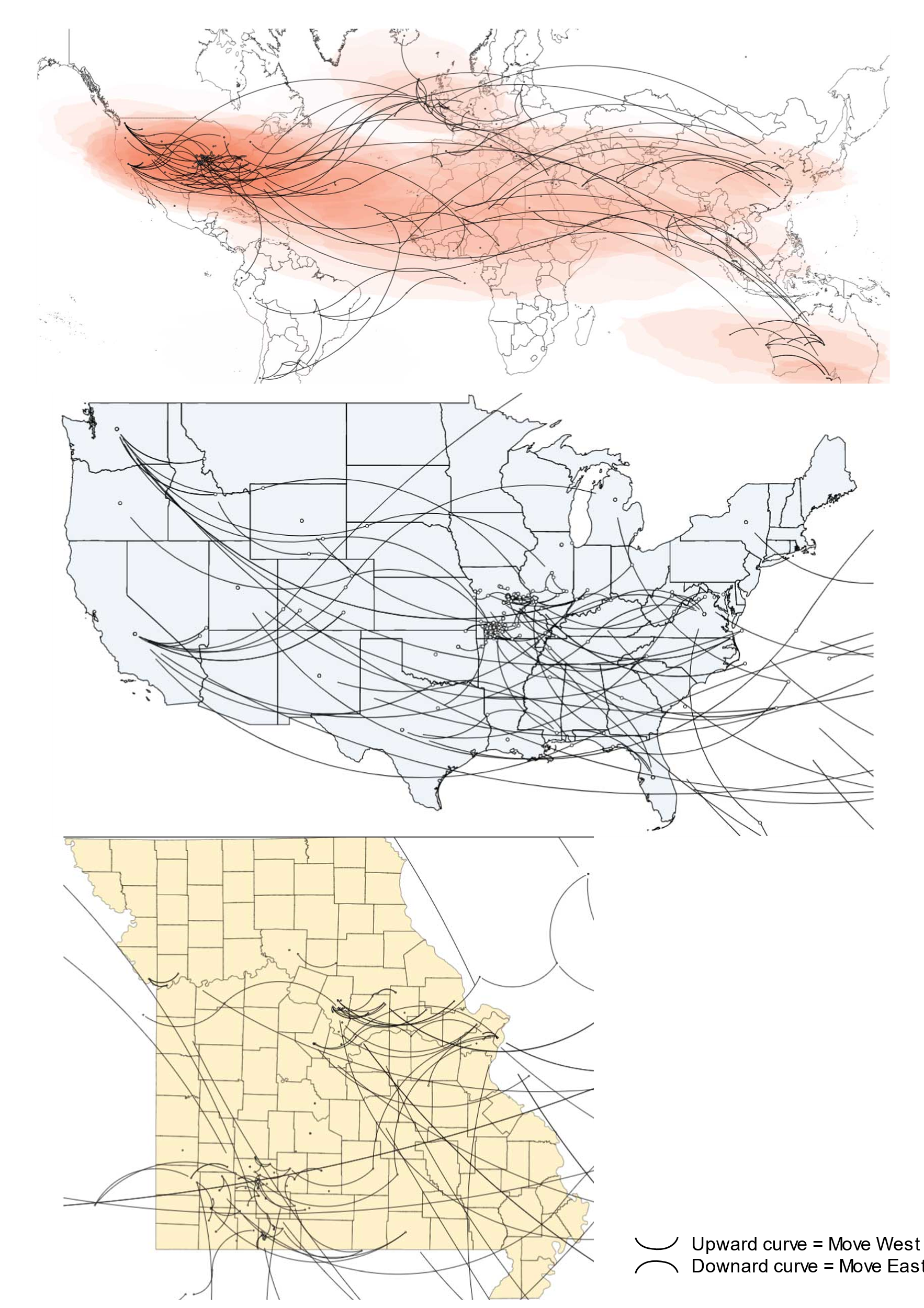

Geospatial distribution and migration of SARS-CoV-2 in Missouri at A) global, B) national, and C) state level. Phylogeographic visualizations created with SpreaD3 and edited in Adobe Illustrator^13^. Westward movements are depicted by an upward curvature. Eastward movements are depicted by a downward curvature.

**eFigure 4. A case of reinfection**.

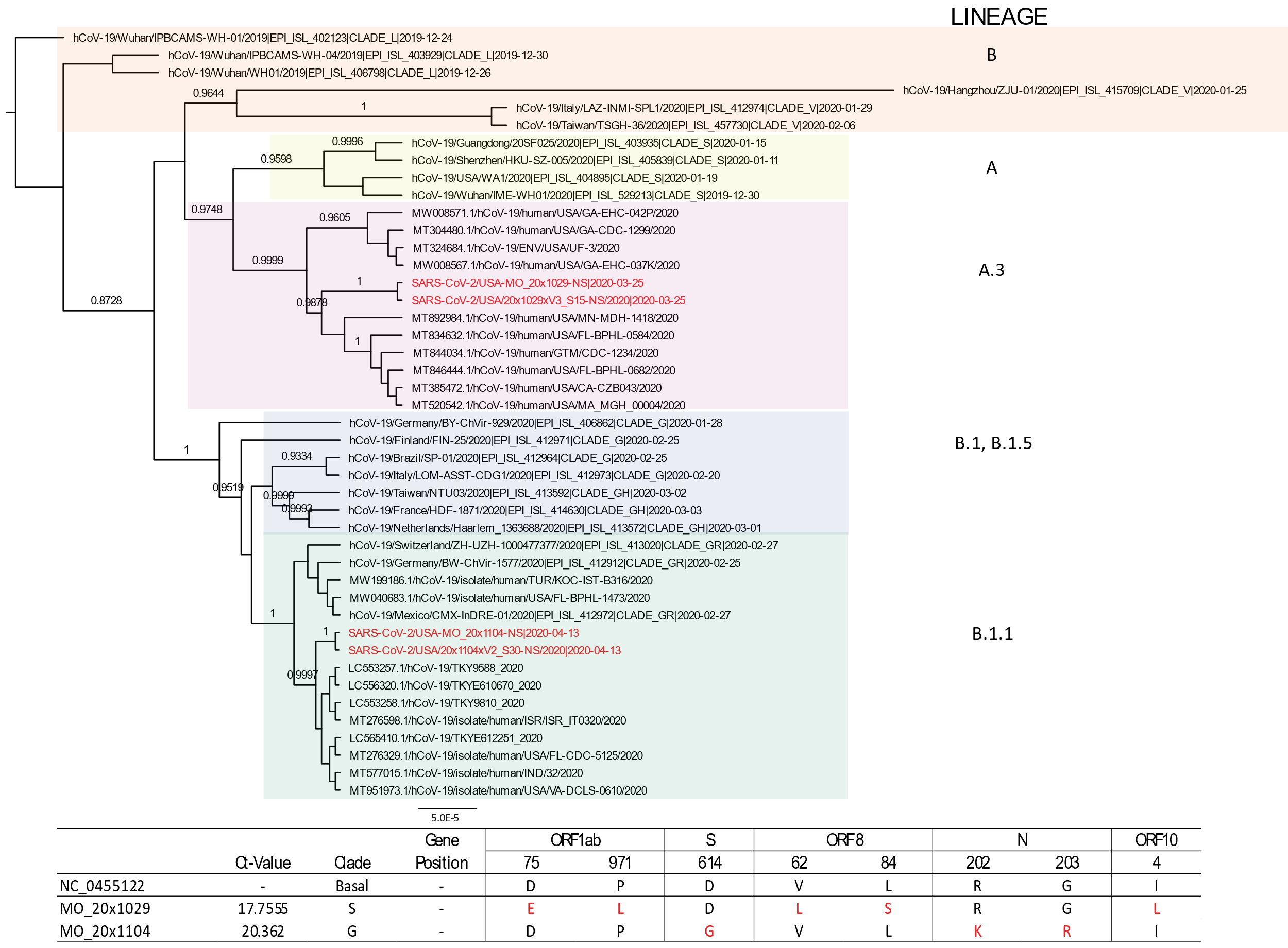

A) Bayesian tree of SARS-CoV-2 viruses with a reinfection case in our study rooted to hCoV-19/Wuhan/PBCAMS-WH-01/2019 (EPI ISL 402123). B) Comparison of specific amino acid mutations of the patient’s 2 strains shown in (B) compared with the reference genome (NC_045512.2). The nomenclature of genetic clades was adapted from the PANGOLIN software. Annotated with taxa names and posterior probabilities > 0.70.

**References**

1.	Li T, Chung HK, Pireku PK, et al. Rapid High Throughput Whole Genome Sequencing of SARS-CoV-2 by using One-step RT-PCR Amplification with Integrated Microfluidic System and Next-Gen Sequencing. bioRxiv 2020.

2.	Wan XF, Chen G, Luo F, Emch M, Donis R. A quantitative genotype algorithm reflecting H5N1 Avian influenza niches. Bioinformatics 2007;23:2368-75.

3.	Wan XF, Ozden M, Lin G. Ubiquitous reassortments in influenza A viruses. J Bioinform Comput Biol 2008;6:981-99.

4.	Zhao ZM, Shortridge KF, Garcia M, Guan Y, Wan XF. Genotypic diversity of H5N1 highly pathogenic avian influenza viruses. J Gen Virol 2008;89:2182-93.

5.	Wu X, Wan XF, Wu G, Xu D, Lin G. Phylogenetic analysis using complete signature information of whole genomes and clustered Neighbour-Joining method. Int J Bioinform Res Appl 2006;2:219-48.

6.	Wu X, Goebel R, Wan XF, Lin G. Whole genome composition distance for HIV-1 genotyping. Comput Syst Bioinformatics Conf 2006:179-90.

7.	Lin G, Cai Z, Wu J, Wan XF, Xu L, Goebel R. Identifying a few foot-and-mouth disease virus signature nucleotide strings for computational genotyping. BMC Bioinformatics 2008;9:279.

8.	Edgar RC. MUSCLE: multiple sequence alignment with high accuracy and high throughput.

9.	Bouckaert R, Heled J, Kühnert D, et al. BEAST 2: a software platform for Bayesian evolutionary analysis. PLoS Comput Biol 2014;10:e1003537.

10.	Kumar S, Stecher G, Li M, Knyaz C, Tamura K. MEGA X: Molecular Evolutionary Genetics Analysis across Computing Platforms. Molecular Biology and Evolution 2018;35:1547-9.

11.	Yang Z. PAML 4: Phylogenetic Analysis by Maximum Likelihood. Molecular Biology and Evolution 2007;24:1586-91.

12.	Lole KS, Bollinger RC, Paranjape RS, et al. Full-length human immunodeficiency virus type 1 genomes from subtype C-infected seroconverters in India, with evidence of intersubtype recombination. J Virol 1999;73:152-60.

13.	Bielejec F, Baele G, Vrancken B, Suchard MA, Rambaut A, Lemey P. SpreaD3: Interactive Visualization of Spatiotemporal History and Trait Evolutionary Processes. Mol Biol Evol 2016;33:2167-9.
